# Analysis of screening results of high-risk population of stroke in Northern Henan Province

**DOI:** 10.1101/2022.01.17.22269405

**Authors:** Zhenhua Li, Qingcheng Yang, Xiangdong Zhang, Yanping Guo, Jiangang Zhang, Ke Sun, Junzheng Yang

## Abstract

**Objectives:** Through the screening analysis of stroke population in Northern Henan from 2014 to 2020, to understand the distribution characteristics of high-risk population of stroke and the correlation between high-risk population of stroke and risk factors, to provide basis for stroke prevention and treatment in Northern Henan province.

**Methods:** A random cluster sampling method was used for screening the high-risk population of stroke in Northern Henan Province, and a questionnaire survey, physical examination and laboratory examination were conducted on the respondents (those who lived in the local area for half a year or more) aged above 40 years old who voluntarily participated in and signed the informed consent, and then the data were sorted and analyzed.

**Results:** A total of 16815 people were participated in screening of high-risk population of stroke in Northern Henan Province, including 6366 males and 10448 females, accounting for 37.86% and 62.14%, respectively; 8855 people were urban household registration, and 7960 people were rural household registration; the age range was 40-80 years old; statistical results demonstrated that there were 3428 people at high-risk of stroke, 3629 people were at moderate risk of stroke, and 8428 people were at low-risk of stroke, accounting for 20.39%, 21.58% and 50.01% in the total screening people, respectively; there were 365 transient Islamic attacks (TIA) patients and 965 stroke patients in the total screening participants in Northern Henan province from 2014 to 2020, accounting for 2.17% and 5.74% in the total screening people, respectively.

After further data sorting, we obtained 2588 valid data of high-risk of stroke, there were significant differences in gender, household registration type and age (P<0.01); the incidence rate of high-risk population of stroke was related to dyslipidemia, atrial fibrillation, smoking, lack of exercise, overweight, family history of stroke, hypertension and diabetes mellitus (P<0.01); the top three highest exposure rates of risk factors were diabetes mellitus (67.16%), overweight (62.60%), and lack of exercise (58.27%) in the high-risk population of stroke. A total of 606 valid data of stroke were screened out, there were significant differences in gender and age (P<0.01); the incidence rate of stroke was related to dyslipidemia, smoking, lack of exercise, overweight, stroke family history, hypertension and diabetes mellitus (P<0.01), the top three highest exposure rates of risk factors of stroke were hypertension (73.27%), overweight (42.08%) and lack of exercise (33.83%) in the stroke patients.

**Conclusion:** The incidence rate of stroke high-risk population and stroke patients was lower in Northern Henan Province compared with national average incidence rate in China, and there are statistical differences at gender and age for the incidence rate of stroke high-risk population and stroke patients in Northern Henan Province. There was the highly correlation between risk factors (dyslipidemia, atrial fibrillation, smoking, lack of exercise, overweight, stroke family history, hypertension and diabetes) and stroke high-risk population or stroke. Regular population screening and early prevention should be taken to reduce the incidence rate of stroke.

## Introduction

Stroke is becoming a kind of global diseases with high incidence rate and high mortality rate in both developing and developed countries, Stroke has been ranked as the second leading cause of death after cancer, it is estimated that stroke has killed at least 6 million 500 thousand people in the world[1]. In China, stroke is also a serious threat to people’s life, there are about 2 million new stroke patients every year, and 7 million now suffer from stroke,which will seriously affect people’s health and cause a great economic burden [2, 3, 4]. Studies have found that several risk factors that influence the development of stroke, such as smoking and hypertension, not only results in the huge economic burden of stroke patients, but also are the main causes of high incidence rate of stroke. In recent years, many evidences demonstrated that early intervention of these risk factors can significantly reduce the incidence of stroke [5, 6]. Therefore, early detection and prevention of stroke are crucial to the treatment of stroke and the reduction of stroke incidence rate. For this purpose, we investigated and analyzed the distribution characteristics of high-risk population of stroke and the correlation between high-risk population of stroke and risk factors in Northern Henan province, which may provide basis for stroke prevention and treatment.

## 1 Subjects and methods

### 1.1 Subjects

Permanent residents over 40 years old in Northern Henan province (those who lived in the local area for half a year or more).

### 1.2 Methods

Using cluster sampling method, the permanent residents over 40 years old in Northern Henan were investigated, mainly including the basic information, lifestyle (smoking, drinking, bad dietary habits, and lack of exercise), family history (whether there is a history of stroke, coronary heart disease, hypertension, diabetes, whether there are stroke, heart disease, hypertension, diabetes and dyslipidemia, and the treatment, control and change of the subjects). Physical examination (including height, weight, waist measurement and blood pressure) and laboratory examination.

And the resident population questionnaire and the informed consent was signed.

### 1.3 Classification of stroke risk assessment

#### (1) 8+2 stroke risk factors and criteria

1. High blood pressure. It should meet any of the following requirements:
  a. History of hypertension (diagnosed by second-level hospital or above);
  b. systolic blood pressure⩾140mmHg or diastolic pressure⩾90mmHg.
2. Atrial fibrillation or valvular heart disease. It should meet any of the following requirements:
  a. Previous medical history (diagnosed by second-level hospital or above).
  b. The results of ECG showed atrial fibrillation.
3. Smoking. Smokers: those who smoke continuously or cumulatively for 6 months or more in their life. Non-smokers: Smokers were quitting smoking at the time of the survey and persist for more than 6 months.
4. Dyslipidemia. It should meet any of the following requirements:
  a. Previous medical history (diagnosed by second-level hospital or above).
  b. Total cholester ⩾ 6.22 mmol/L (240mg/dl), Triglycerid ⩾ 2.3mmol/l (200mg/dl), HDL<1.04 mmol/L (40mg/dl), LDL⩾4.1 mmol/L(160 mg/dl), of which one or more abnormalities can be determined as dyslipidemia.
5. Diabetes mellitus. It should meet any of the following requirements:
  a. Previous medical history (diagnosed by second-level hospital or above).
  b. The random blood glucose ⩾ 11.0 mmol/L or fasting blood glucose ⩾ 7.0mmol/L.
6. Little physical exercise. Exercise⩾3 times a week, moderate intensity and above exercise⩾30 minutes each time, or engaged in moderate and severe physical workers are regarded as regular physical exercise. On the contrary, it is lack of exercise ; moderate physical labor refers to the continuous movement of hands and arms (such as sawing wood); work of arms and legs (such as transport operations of trucks, tractors or construction equipment); arm and trunk work (such as forging, pneumatic tool operation, painting, intermittent handling of medium weight, weeding, hoeing, picking fruits and vegetables, etc.).Heavy physical labor refers to the work of arm and trunk load (such as lifting heavy objects, shoveling, hammering, sawing or chiseling hardwood, mowing, excavation, etc.).
7. Obesity. BMI⩾28 is obesity. [(BMI=weight (kg)/height^2^ (m^2^)].
8. Family history of stroke (diagnosed by second-level hospital or above).
  a. Previous stroke history (diagnosed by CT in second-level hospital or above).
  b. TIA history (diagnosed by CT in second-level hospital or above).

#### (2) Classification of high-risk population of stroke, medium-risk population of stroke and low-risk population of stroke

High risk population of stroke: there are 3 or 3 above of the 8 risk factors (hypertension, dyslipidemia, diabetes, atrial fibrillation or valvular heart disease, smoking history, obesity, lack of exercise, family history of stroke), or transient ischemic attack, or previous stroke patients were diagnosed in high-risk population of stroke.

Medium-risk population of stroke: There are less than 3 of the 8 risk factors (hypertension, dyslipidemia, diabetes, atrial fibrillation or valvular heart disease, smoking history, obesity, lack of exercise, family history of stroke), but including one of the three chronic diseases (hypertension, diabetes, atrial fibrillation or valvular heart disease).

Low-risk population of stroke: there are less than 3 of the 8 risk factors (hypertension, dyslipidemia, diabetes, atrial fibrillation or valvular heart disease, smoking history, obesity, lack of exercise, family history of stroke), but not including one of the three chronic diseases (hypertension, diabetes, atrial fibrillation or valvular heart disease).

### 1.4 Statistical analysis

Data sorting was used by Excel software, SPSS 22.0 was used for statistical analysis, and the measurement data are described as X±s; The counting data are described by percentage (%) and compared by χ^2^ test, and P<0.05 was considered statistically significant.

## 2 Results

### 2.1 The basic information of the screened participants in Northern Henan province from 2014 to 2020

Firstly, Annual screening results demonstrated that the basic information of the participants including gender, Household registration type and age distribution who participated in stroke screening activity in Northern Henan province from 2014 to 2020 every year (Table 1). And then, totally analysis showed that there are 16815 people were participated in stroke screening activity in Northern Henan province from 2014 to 2020, including 6367 males and 10448 females; 8855 people were urban household registration, and 7960 people were rural household registration; The age of screening participants were 40-80 years old (Table 2).

**Table 1.**
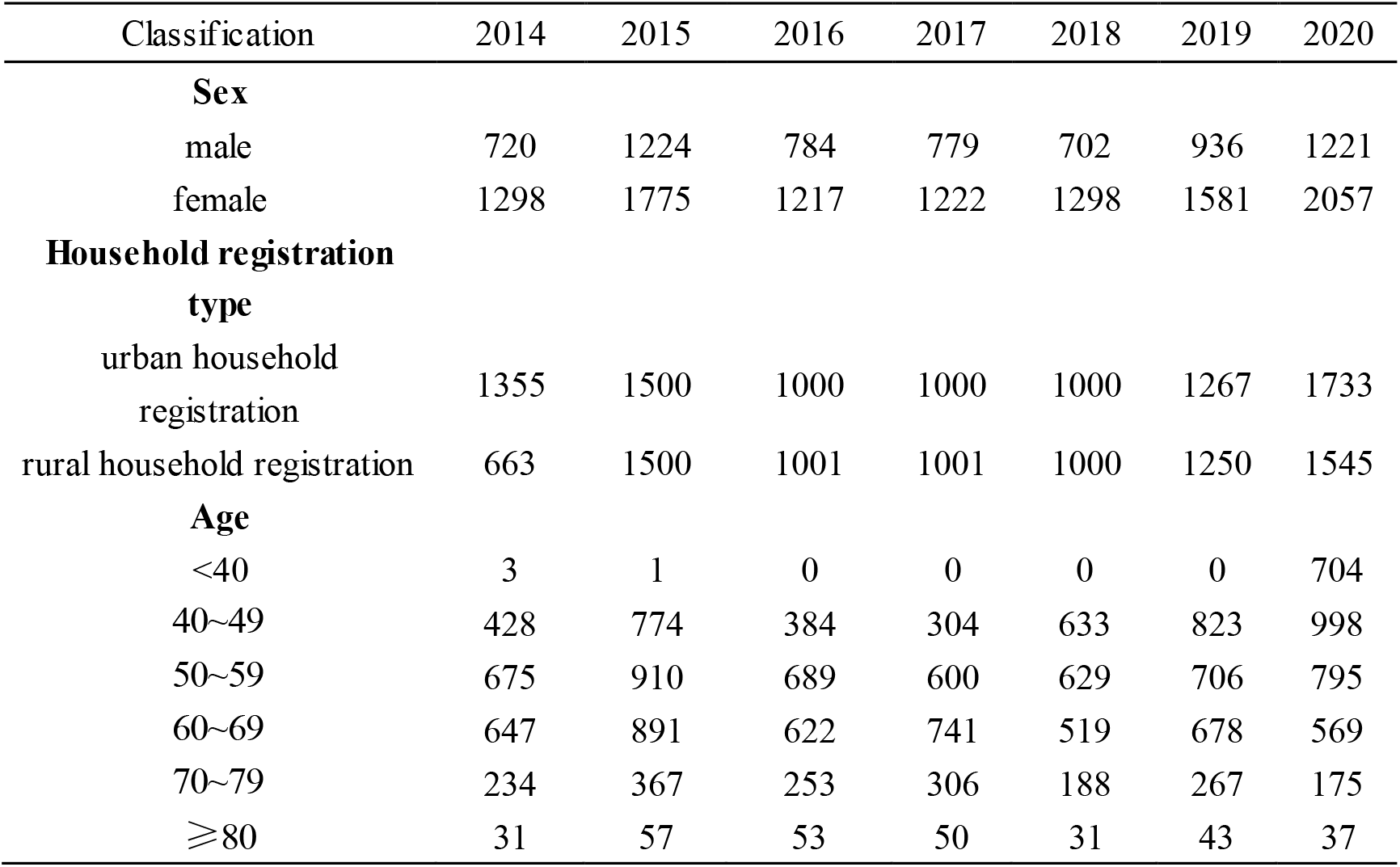
Annual screening of basic information of people who participated in stroke screening activity in Northern Henan province from 2014 to 2020.

**Table 2.** Comprehensive analysis of basic information of people who participated in stroke screening activity in Northern Henan province from 2014 to 2020.

| Classification | Number | proportion/% |
| --- | --- | --- |
| **Sex** |  |  |
| male | 6367 | 37.86% |
| female | 10448 | 62.14% |
| **Household registration type** |  |  |
| urban household registration | 8855 | 52.66% |
| rural household registration | 7960 | 47.34% |
| **Age** |  |  |
| <40 | 708 | 4.21% |
| 40~49 | 4344 | 25.83% |
| 50~59 | 5004 | 29.76% |
| 60~69 | 4667 | 27.75% |
| 70~79 | 1790 | 10.65% |
| ≥80 | 302 | 1.80% |

**Table 3.**
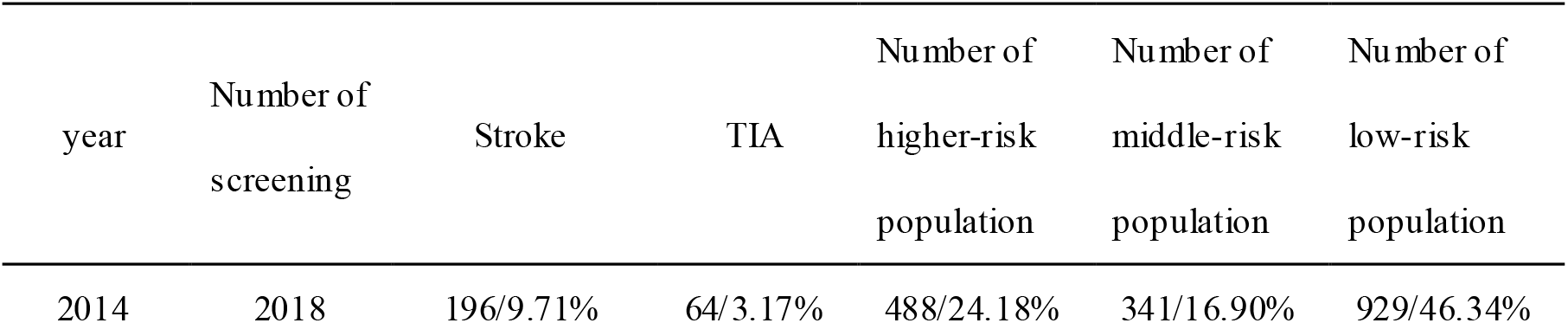
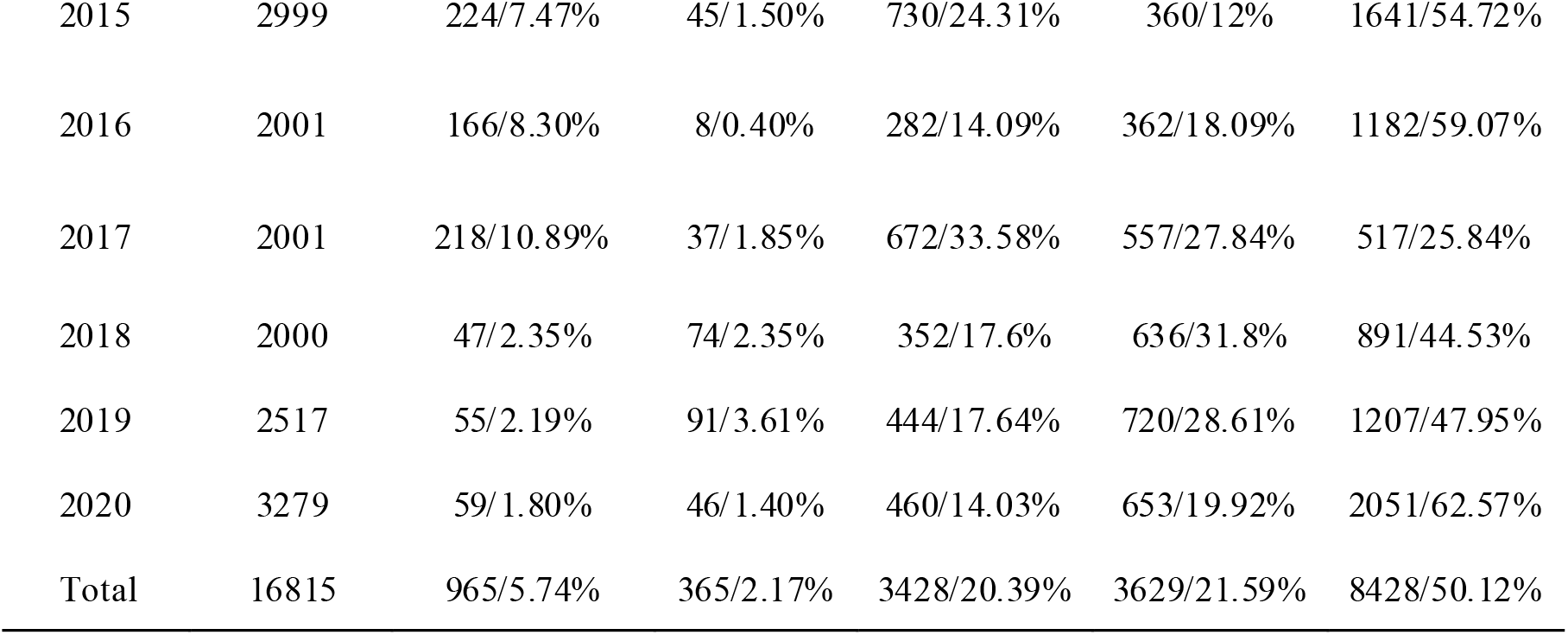
The distribution of stroke, TIA and high-risk population of stroke in Northern Henan province every year from 2014 to 2020.

**Table 3.**
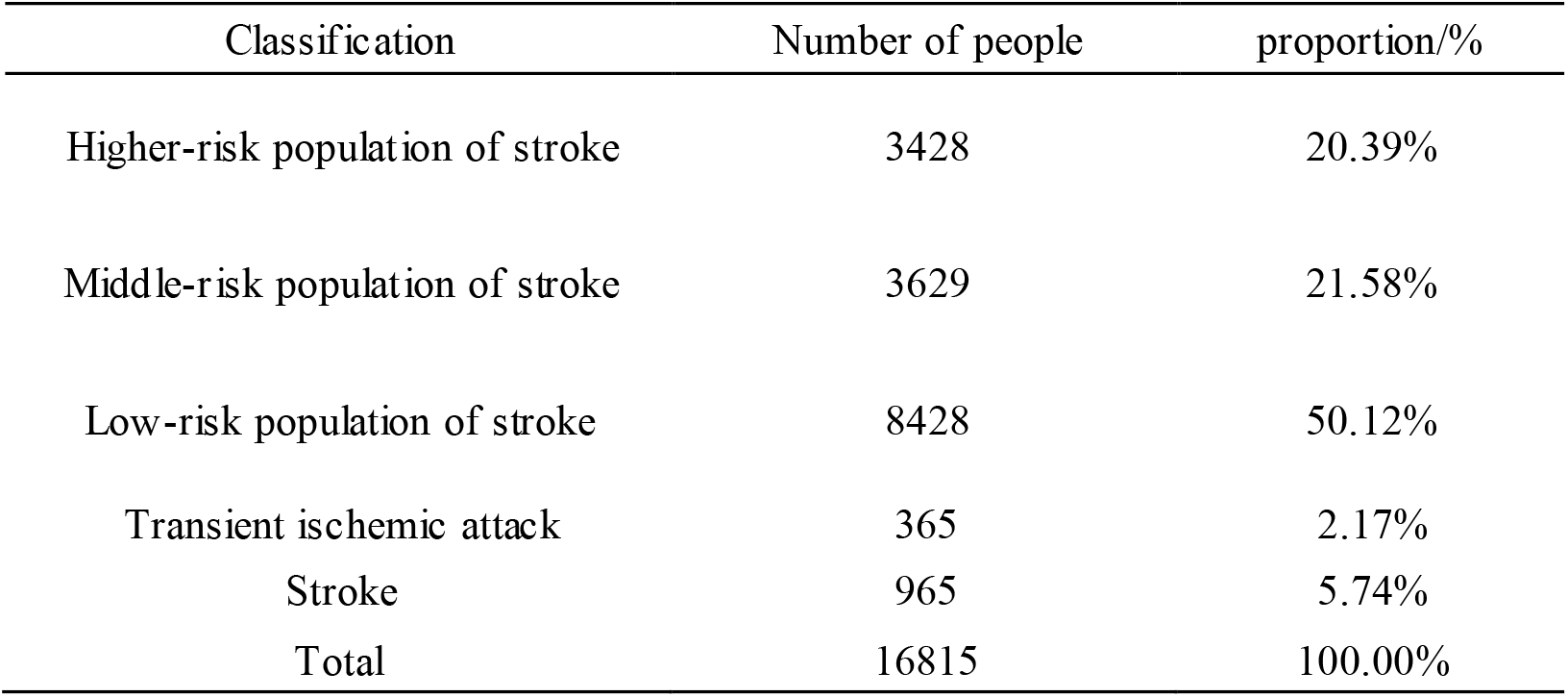
The overall distribution of stroke, TIA and high-risk population of stroke in Northern Henan province from 2014 to 2020

**Table 4.** The distribution of stroke high-risk population of stroke in Northern Henan province from 2014 to 2020

| Classification | Total | High-risk<br>population of<br>stroke | Proportion /% | $\chi^2$ | $P$ |
| --- | --- | --- | --- | --- | --- |
| **Sex** |  |  |  |  |  |
| Male | 6366 | 1301 | 20.44% | 200.208 | <0.01 |
| Female | 10448 | 1287 | 12.32% |  |  |
| **Household**<br>**registration type** |  |  |  |  |  |
| urban household<br>registration | 8855 | 1799 | 20.32% | 348.445 | <0.01 |
| rural household<br>registration | 7960 | 789 | 9.91% |  |  |
| **Age** |  |  |  |  |  |
| <40 | 708 | 1 | 0.14% | 33.741 | <0.01 |
| 40~49 | 4344 | 578 | 13.31% |  |  |
| 50~59 | 5004 | 858 | 17.15% |  |  |
| 60~69 | 4667 | 790 | 16.93% |  |  |
| 70~79 | 1790 | 333 | 18.60% |  |  |
| ≥80 | 302 | 28 | 9.27% |  |  |

**Table 5.** The distribution of risk factors of stroke high-risk population of stroke in Northern Henan province from 2014 to 2020

| Risk factor | High-risk<br>population of<br>stroke | Proportion/% | $\chi^2$ | *P* |
| --- | --- | --- | --- | --- |
| Dyslipidemia |  |  |  |  |
| Yes | 1251 | 0.4834 | 1533.876 | <0.01 |
| No | 1337 | 0.5166 |  |  |
| Atrial fibrillation |  |  |  |  |
| Yes | 101 | 0.039 | 247.984 | <0.01 |
| No | 2487 | 0.961 |  |  |
| Smoking |  |  |  |  |
| Yes | 923 | 0.3566 | 621.902 | <0.01 |
| No | 1665 | 0.6434 |  |  |
| Lack of exercise |  |  |  |  |
| Yes | 1508 | 0.5827 | 2475.762 | <0.01 |
| No | 1080 | 0.4173 |  |  |
| Overweight |  |  |  |  |
| Yes | 1620 | 0.626 | 2942.414 | <0.01 |
| No | 968 | 0.374 |  |  |
| Family history of stroke |  |  |  |  |
| Yes | 1304 | 0.5039 | 1712.757 | <0.01 |
| No | 1284 | 0.4961 |  |  |
| Hypertension |  |  |  |  |
| Yes | 638 | 0.2465 | 138.738 | <0.01 |
| No | 1950 | 0.7535 |  |  |
| Diabetes mellitus |  |  |  |  |
| Yes | 1738 | 0.6716 | 3468.707 | <0.01 |
| No | 850 | 0.3284 |  |  |

**Table 6.** The distribution of stroke patients in Northern Henan province from 2014 to 2020

| Classification | Screened participants | The number of stroke patients | Proportion/% | $\chi^2$ | $P$ |
| --- | --- | --- | --- | --- | --- |
| **Sex** |  |  |  |  |  |
| male | 6366 | 296 | 4.65% | 32.236 | <0.01 |
| female | 10448 | 310 | 2.97% |  |  |
| **Household registration type** |  |  |  |  |  |
| urban household registration | 8855 | 301 | 3.40% | 2.257 | >0.05 |
| rural household registration | 7960 | 305 | 3.83% |  |  |
| **Age** |  |  |  | 284.878 | <0.01 |
| <40 | 708 | 0 | 0.00% |  |  |
| 40~49 | 4344 | 30 | 0.69% |  |  |
| 50~59 | 5004 | 135 | 2.70% |  |  |
| 60~69 | 4667 | 281 | 6.02% |  |  |
| 70~79 | 1790 | 132 | 7.37% |  |  |
| $\geq 80$ | 302 | 28 | 9.27% | | |

**Table 7.** The distribution of risk factors of stroke patients in Northern Henan province from 2014 to 2020

| Risk factor | The number of stroke patients | Proportion/% | $\chi^2$ | $P$ |
| --- | --- | --- | --- | --- |
| Dyslipidemia |  |  |  |  |
| Yes | 197 | 32.51% | 1140.427 | <0.01 |
| No | 409 | 67.49% |  |  |
| Atrial fibrillation |  |  |  |  |
| Yes | 30 | 4.95% | 3.438 | >0.05 |
| No | 576 | 95.05% |  |  |
| Smoking |  |  |  |  |
| Yes | 148 | 24.42% | 629.543 | <0.01 |
| No | 458 | 75.58% |  |  |
| Lack of exercise |  |  |  |  |
| Yes | 205 | 33.83% | 1234.975 | <0.01 |
| No | 401 | 66.17% |  |  |
| Overweight |  |  |  |  |
| Yes | 255 | 42.08% | 1888.568 | <0.01 |
| No | 351 | 57.92% |  |  |
| Family history of stroke |  |  |  |  |
| Yes | 201 | 33.17% | 1187.334 | <0.01 |
| No | 405 | 66.83% |  |  |
| Hypertension |  |  |  |  |
| Yes | 444 | 73.27% | 5125.032 | <0.01 |
| No | 162 | 26.73% |  |  |
| Diabetes mellitus |  |  |  |  |
| Yes | 145 | 23.93% | 602.357 | <0.01 |
| No | 461 | 76.07% |  |  |

### 2.2 The screening results of stroke, TIA and high-risk population of stroke in Northern Henan province from 2014 to 2020

There were 965 stroke patients were screened in Northern Henan province from 2014 to 2020, accounting for 5.74% of the total screening participants; 365 cases of TIA were screened, accounting for 2.17% of the total screening participants; 3428 people were at high-risk population of stroke, accounting for 20.39% of the total screening participants; 3629 people were at moderate-risk population of stroke, accounting for 21.58% of the total screening participants; 8428 people were at low-risk population of stroke, accounting for 50.01% of the total screening participants (Table 4).

### 2.3 The distribution of high-risk population of stroke in Northern Henan province from 2014 to 2020

The basic information of stroke high-risk groups in Northern Henan province from 2014 to 2020 was sorted out according to gender, Household registration type and age. there were 1301 males in stroke high-risk population of stroke in Northern Henan province from 2014 to 2020, accounting for 20.44% of the total screening male participants, there were 1287 females in stroke high-risk population of stroke in Northern Henan province from 2014 to 2020, accounting for 12.32% of the total screening female participants, there was statistical significance between the two group (P<0.01); there were 1799 urban household registration, accounting for 20.32% of the high-risk population of stroke with urban household registration, and there were 789 rural household registration, accounting for 9.91% of the high-risk population of stroke with rural household registration (P<0.01); The age of high-risk population of stroke was 40-80 years old, and the age distribution of high-risk population of stroke in Northern Henan from 2014 to 2020 was statistically significant (P<0.01).

The risk factors related to the 2588 high-risk stroke patients include dyslipidemia, atrial fibrillation, smoking, and lack of exercise, overweight, family history of stroke, hypertension and diabetes. The top three exposure rates were diabetes Mellitus (67.16%), overweight (62.60%), and lack of exercise (58.27%). Dyslipidemia, atrial fibrillation, smoking, lack of exercise, overweight, family history of stroke, hypertension and diabetes mellitus were all high closely related to stroke, there was a statistical significance in the distribution of each risk factor (P<0.01).

### 2.3 The distribution of stroke patients in Northern Henan province from 2014 to 2020

Then, the basic information of stroke population in Northern Henan province from 2014 to 2020 was sorted out according to gender, household registration type and age. It was found that there were 296 male stroke patients in Northern Henan province from 2014 to 2020, accounting for 4.65% of the total screening male participants, and there were 310 female stroke patients in Northern Henan province from 2014 to 2020, accounting for 2.97% of the total screening female participants. There was statistical significance between two group (P<0.01); there were 301 urban household registration, accounting for 3.40% of the total stroke patients with urban household registration, and there were 305 rural household registration, accounting for 3.83% of the total stroke patients with rural household registration, there was no significant difference between two kinds of household registration types (P>0.05); the age of stroke patients were 40-80 years old, and there was statistically significant at the age distribution of stroke patients (P< 0.01).

Among the 2588 stroke patients, the top three highest risk factor exposure rates were hypertension (73.27%), overweight (42.08%) and lack of exercise (33.83%); dyslipidemia, atrial fibrillation, smoking, lack of exercise, overweight, family history of stroke, hypertension and diabetes were highly related to stroke, there was a statistical significance in the distribution of each risk factor for stroke patients except Atrial fibrillation (P<0.01).

## 3 Discussion

With the development of economy, the number of stroke patients in China is increasing, which has caused a huge economic burden to the family and the country. Therefore, it is of great clinical significance to timely screen and prevent the occurrence and development of stroke.

In this study, we summarized the stroke screening data in Northern Henan province from 2014 to 2020, summarized the stroke screening results according to age, household registration type and age, stroke high-risk population, medium risk population, low-risk population and stroke risk factors, and found that there were 2588 stroke high-risk population of stroke in Northern Henan province from 2014 to 2020, accounting for 15.39% in the total screening population; it is lower than 18.51% (reported in China stroke prevention and control report 2018) and 19.84% (reported in China stroke prevention and control report 2019), indicating that the stroke situation in Northern Henan province was not very severe; female high-risk population of stroke in Northern Henan from 2014 to 2020 was slightly higher than male, and there was significant difference between male and female of high-risk population of stroke in Northern Henan from 2014 to 2020; the high-risk population of stroke from urban household registration is significantly higher than that from rural household registration; the incidence rate of stroke in North Henan is 5.74%, and there are statistical differences between male and female gender, household type and age. These data suggest that the distribution of high-risk population of stroke in Northern Henan province is relatively complex.

The top three risk factors of high-risk population of stroke were diabetes mellitus (67.16%), overweight (62.60%) and lack of exercise (58.27%); the top three risk factors of stroke of exposure rates were hypertension (73.27%), overweight (42.08%) and lack of exercise (33.83%); those evidences demonstrated that overweight and lack of exercise maybe the main risk factor in Northern Henan province.

Based on the above, the distribution of stroke high-risk groups and stroke patients in Northern Henan province is very characteristic, and the necessary precautions need to be taken.

## Data Availability

I have the right to post this manuscript and confirm that all authors have assented to posting of the manuscript. All relevant ethical guidelines have been followed. Details of the oversight body: Ethics committee of Anyang People's Hospital suggested that Analysis of screening results of high-risk population of stroke in Northern Henan Province is ethical in according with Ethics committee of Anyang People's Hospital, and agree to conduct relevant experiments. All necessary patient/participant consent has been obtained. I am legally responsible for the content of the article. I have followed all appropriate research reporting guidelines.
All necessary patient/participant consent has been obtained and the appropriate institutional forms have been archived.

## 5 Acknowledgments

None.

